# Cardiovascular Risks Associated with Patients Infected with Scrub Typhus: A Nationwide Cohort Study

**DOI:** 10.1101/2023.07.17.23292799

**Authors:** Jih-Kai Yeh, Victor Chien-Chia Wu, Shao-Wei Chen, Chia-Ling Wu, Yu-Sheng Lin, Chun-Wen Cheng, Chih-Hsiang Chang, Michael Wu, Pao-Hsien Chu, Shang-Hung Chang, Yu-Tung Huang

**Author notes:** Address for Co-correspondence: Yu-Tung Huang, PhD Center for Big Data Analytics and Statistics, Chang Gung Memorial Hospital, Linkou Medical Center No. 5, Fuxing Street, Guishan District, Taoyuan City 33305, Taiwan.

## Abstract

**Background:** Scrub typhus is an infectious disease that affects multiple organs. However, the long-term cardiovascular (CV) risk in survivors remains unknown.

**Method:** A retrospective cohort study used administrative claims data from the National Health Insurance Research Database (NHIRD) to investigate the CV risk of scrub typhus survivors from January 1, 2010, to December 31, 2015. People who had prior CV events before the diagnosis of scrub typhus were excluded. The CV outcomes of interest were acute myocardial infarction (AMI), heart failure hospitalization (HFH), hemorrhagic or ischemic stroke, new-onset atrial fibrillation (AF), aneurysm or dissection of aorta, venous thromboembolism (VTE), and CV death.

**Result:** A total of 2,269 patients with scrub typhus and without a prior CV event were identified (mean age 47.8±16.1 years, 38.0% female). The health control cohort (n=2,264) was selected to compare by the frequency matching with age, gender, and co-morbidities with patients with scrub typhus. The incidence of HFH, new-onset AF, and total events was significantly higher among patients with scrub typhus than the control cohort with an adjusted hazard ratio (aHR) of 1.97, 95% confidence interval (CI) 1.13-3.42 for HFH; 2.48, 95% CI: 1.23-5.0 for new-onset AF; 1.43, 95% CI: 1.08-1.91 for total CV events, respectively. The event rates of other outcomes were similar between the two groups.

**Conclusion:** In the cohort study, survivors of scrub typhus are at heightened risk of subsequent CV events, especially for HFH and new-onset AF. These findings serve as an important reminder to physicians regarding the significant CV risk that remains present following acute scrub typhus infection.

## Background

Scrub typhus is an infection caused by Orientia tsutsugamushi, a mite-borne rickettsia. The disease is typically endemic in Asia-Pacific regions, but there have been increasing reports of outbreaks in non-endemic areas such as Africa, France, the Middle East, and South America^1, 2^. The clinical presentation of patients with scrub typhus can vary from a nonspecific flu-like illness to life-threatening organ dysfunction, septic shock, and death^3, 4^. Severe scrub typhus is more likely to occur in the elderly, those with a longer duration of illness (>7 days), and those without the characteristic eschar^5^. While the presence of an eschar, which typically develops in covered areas of the skin such as the axilla and groin, can be a clue for early diagnosis, it is only present in 40-55% of patients^6, 7^. The diagnosis of scrub typhus still relies on a high index clinical suspicion, especially in patients with history of exposure in the endemic region.

The principal pathophysiologic finding of scrub typhus is widespread vasculitis, perivascular inflammation, altered capillary permeability, and microangiopathy in the involved organs^8^. Scrub typhus infection with cardiac involvement is common and have been reported in prospective cohort studies, with reduced left ventricular ejection fraction in 31-43% of patients; elevated cardiac troponin T levels in 62-73% of patients^9, 10^. In addition, the abnormal electrocardiogram (ECG) finding of ST-segment changes and T wave inversion were also observed in 9.8-66% and 8.5-12.3% of patients, respectively^9, 10^. The degree of prolonged QT interval was found to be associated with the severity of scrub typhus infection^11^. These ECG changes are thought to be related to the active inflammation in the myocardium and are correlated to the area and degree of myocardial injury during infectious disorders^12^. Although patients with milder disease typically recover within 48 hours of initiating appropriate antibiotics, subclinical structural and functional impairment in the cardiovascular (CV) system may go unrecognized. Currently, comprehensive study on the subsequent development of cardiovascular diseases in patients with scrub typhus infection is lacking. Therefore, we conducted a longitudinal nationwide cohort study utilizing the Notifiable Infectious Diseases (NID) database from Taiwan Centers for Disease Control (Taiwan CDC) with linkage to the National Health Insurance (NHI) claims data. The objective of the study was to investigate whether patients with scrub typhus have an increased risk for acute myocardial infarction (AMI), hospitalization for heart failure (HFH), new-onset atrial fibrillation (AF), ischemic or hemorrhagic stroke, common aortic diseases, venous thromboembolism (VTE), and CV death.

## Methods

### Data Source

The data of the study were retrieved from the Taiwan National Health Insurance Research Database (NHIRD) from January 1, 2010, to December 31, 2015. The NHIRD contains all records of inpatient, outpatient, and emergency services with diagnoses, prescriptions, examinations, operations, and expenditures from medical care providers. The database included more than 99% of 23 million Taiwanese population. Taiwan CDC establishes, manages, and keeps track of infectious disease under the NID surveillance reporting system. All cases of scrub typhus were confirmed by the Centers for Disease Control in term of the pathogen isolation and or serological assays. The data linkages between the NHIRD and NID were available at Health and Welfare Data Center (HWDC). All NHIRD and NID databases were encrypted and used for research purpose at national HWDC sites to protect patient privacy.

### Identification of Study Cohort

This is a retrospective nationwide cohort study. All patients with more than 20-year-old and a diagnosis of scrub typhus in NID database between 2010 to 2015 were included (n=2,347). The study index date was defined as the date of the first diagnosis of scrub typhus in outpatient visit or discharge records. Among these patients, we excluded patients who had any of prior major CV events, including AMI, HFH, AF, ischemic or hemorrhagic stroke, aortic aneurysm or dissection, and VTE before the index date (n=78). The comparison group was selected from people under the NIH program who had never been diagnosed with scrub typhus and had no prior history of major CV events before the year of 2010 (n=21,424,000). Each scrub typhus case was matched to one randomly selected control individuals according to age, gender, and comorbidities. Two step frequency matching was used in this study. First, we used 1:20 ratio to select downsizing’s control group by cases’ birth year randomly and obtain the index date for control group. After then, we used one to one ratio matching by birth year, sex, and comorbid conditions, including diabetes mellitus, hypertension, hyperlipidemia, coronary artery disease (CAD), chronic obstructive pulmonary disease (COPD), chronic kidney disease (CKD), chronic liver disease, autoimmune disease, and cancer. Finally, 2,264 patients with scrub typhus and no prior major CV events and 2,264 matched control individuals were identified. The flow chart of study cohort identification was depicted in Figure1.

**Figure 1.**
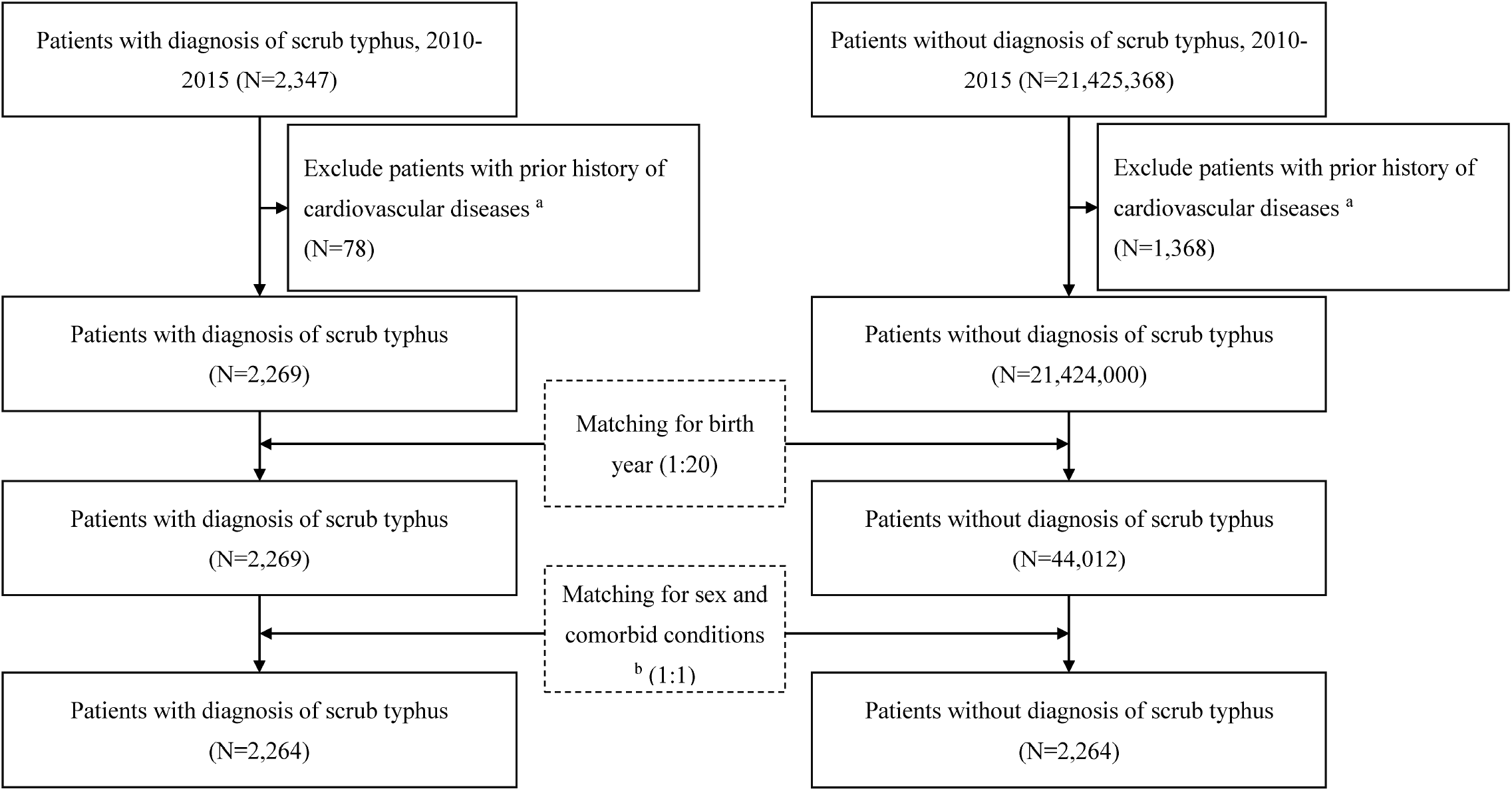
Flow chart of selection and inclusion of patients with scrub typhus and the control population

### Measurement of Covariates and Study Outcomes

The common risk factors of CV diseases were considered as the covariates of this study, such as age, gender, diabetes mellitus, hypertension, hyperlipidemia, CAD, COPD, CKD, chronic liver disease, autoimmune disease, and cancer. Diseases were identified by International Classification of Diseases, 9th Revision, Clinical Modification (ICD-9-CM) codes before 2016 or ICD-10-CM codes thereafter, with diagnostic codes in more than 2 times of outpatient visits or one primary discharge diagnosis within the previous year of the index date. The CV outcomes of interest included AMI, HFH, ischemic stroke, hemorrhagic stroke, new-onset AF, aneurysm or dissection of aorta, VTE, and cardiovascular death. All participants were followed from the index date until the first occurrence of above cardiovascular outcomes, death, withdrawal from insurance, or the end of the follow-up period (December 31, 2017), whichever came first. The ICD-9-CM and ICD-10-CM disease codes were provided in the supplementary table 1.

### Statistical analysis

The frequency matching was used to balance the differences of baseline covariates between patients with scrub typhus and the comparison cohort. The balance of covariates before and after matching was assessed by the absolute standardized mean difference (SMD), which a value less than 0.1 was considered balance distribution between groups. Cumulative probabilities of CV events were depicted by the Kaplan-Meier method and were compared between groups with the log-rank tests. To determine whether patients with scrub typhus had increased the risks of CV events, the incidence rate ratio (IRR) and 95% CI for these events per 1,000 person-years was analyzed by Poisson regression. The Cox proportional hazards regression models were used to calculate the hazard ratios (HRs) and 95% confidence interval (CI) of each specified CV events in scrub typhus and comparison cohort. Subgroup analyses for each specific events were assessed by stratification of age group, sex, and comorbid condition. A two-sided P value < 0.05 was statistically significant. No adjustment of multiple testing (multiplicity) was made in this study. The analyses were performed using the SAS Version 9.4 (SAS Institute, Cary, NC).

### Ethics statement

The study protocol was approved by the Institutional Review Board of Chang Gung Medical Foundation (IRB No: 201901517B0). All data converted from the NHI’s claim records were anonymized and were analyzed on-site at HWDC. This study was conducted in accordance with the Declaration of Helsinki and the Declaration of Taipei on ethical considerations regarding health databases by the World Medical Association.

## Results

### Subject characteristics

The study cohort consisted of 2,269 patients with a diagnosis of scrub typhus with a mean age of 47.8±16.1 years, 38.0% of whom were female. For their comorbidities, 7.1% of them had diabetes mellitus; 14.3% of them had hypertension; 2.6% of them had COPD; 0.3% of them had CKD; 0.4% of them had autoimmune disease; and 2.6% of them had cancer. After 1:1 frequency matching with age, gender and comorbidities, the comparison cohort (n=2264) and study cohort (n=2264) were identified. The distribution of age, gender, and comorbidities between two groups was balanced with all absolute SMD < 0.01. Table 1 presented the baseline characteristics of cohorts with and without scrub typhus before and after the 1:1 frequency matching. The mean follow-up duration in the scrub typhus and comparison groups were 4.5 and 4.6 years, respectively. During this period, 138 participates died and we identified 45 patients with AMI, 56 patients with HFH, 27 patients with hemorrhagic stroke, 63 patients with ischemic stroke, 38 patients with new onset atrial fibrillation, 9 patients with aortic aneurysm or dissection, 11 patients with VTE, and 25 patients with CV death. The incidence of pre-specified CV events between two groups during study period was presented in Table 2. Overall, patients with scrub typhus had increased risk of HFH and new-onset AF compared with individuals in control group (3.62 versus 1.84 per 1000 person-years; 2.63 versus 1.06), with rate ratios of 1.92 (95% CI: 1.10-3.34) and 2.39 (95% CI: 1.18-4.83), respectively. With the Cox regression analysis with adjustment for age, gender, and comorbidities, the hazard ratio of HFH and new-onset AF among patients with scrub typhus than those control individuals were 1.97 (95% CI: 1.13-3.42, p=0.016) and 2.48 (95% CI: 1.23-5.0, p=0.011), respectively. The occurrence of AMI, hemorrhagic stroke, ischemic stroke, aneurysm or dissection of aorta, and CV death were not statistically different between two groups. Nonetheless, all adverse CV events, including AMI, HFH, hemorrhagic or ischemic stroke, new-onset AF, aortic aneurysm or dissection, VTE and CV death, occurred more in patients with scrub typhus than in control cohort, with an adjusted hazard ratio of 1.43 (95% CI: 1.08-1.91, p= 0.013). Cumulative incidence of each cardiovascular outcomes by the Kaplan-Meier method were displayed in figure 2 and 3. Moreover, the effects of scrub typhus infection on subsequent CV risk did not modify in the subgroup analysis, which was stratified by age, genders, and comorbidities (Table 3).

**Figure 2.**
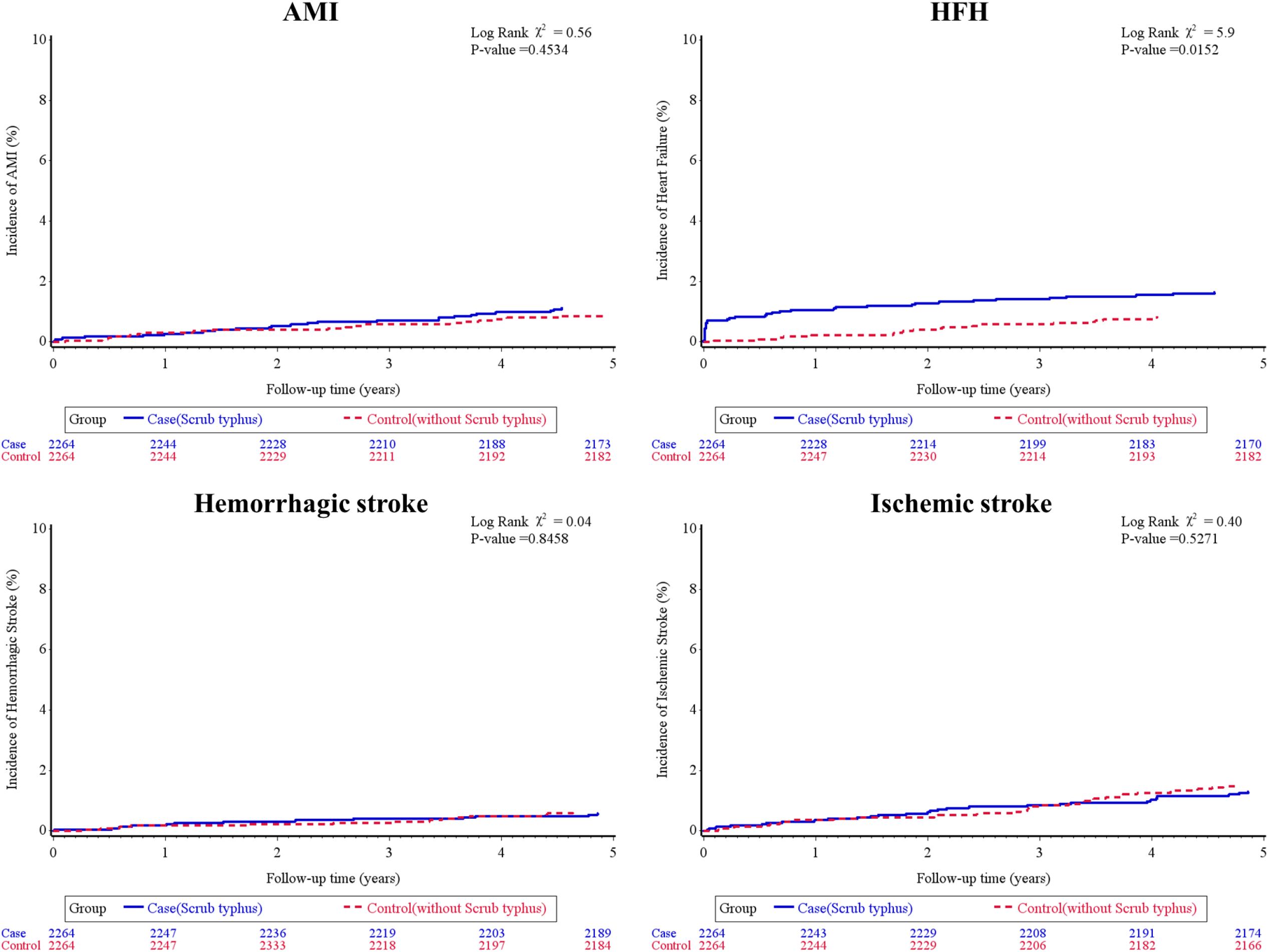
Cumulative incidence of outcome events of acute myocardial infarction (AMI) (A), heart failure hospitalization (HFH) (B), hemorrhagic stroke (C), and ischemic stroke (D) in patients with and without scrub typhus.

**Figure 3.**
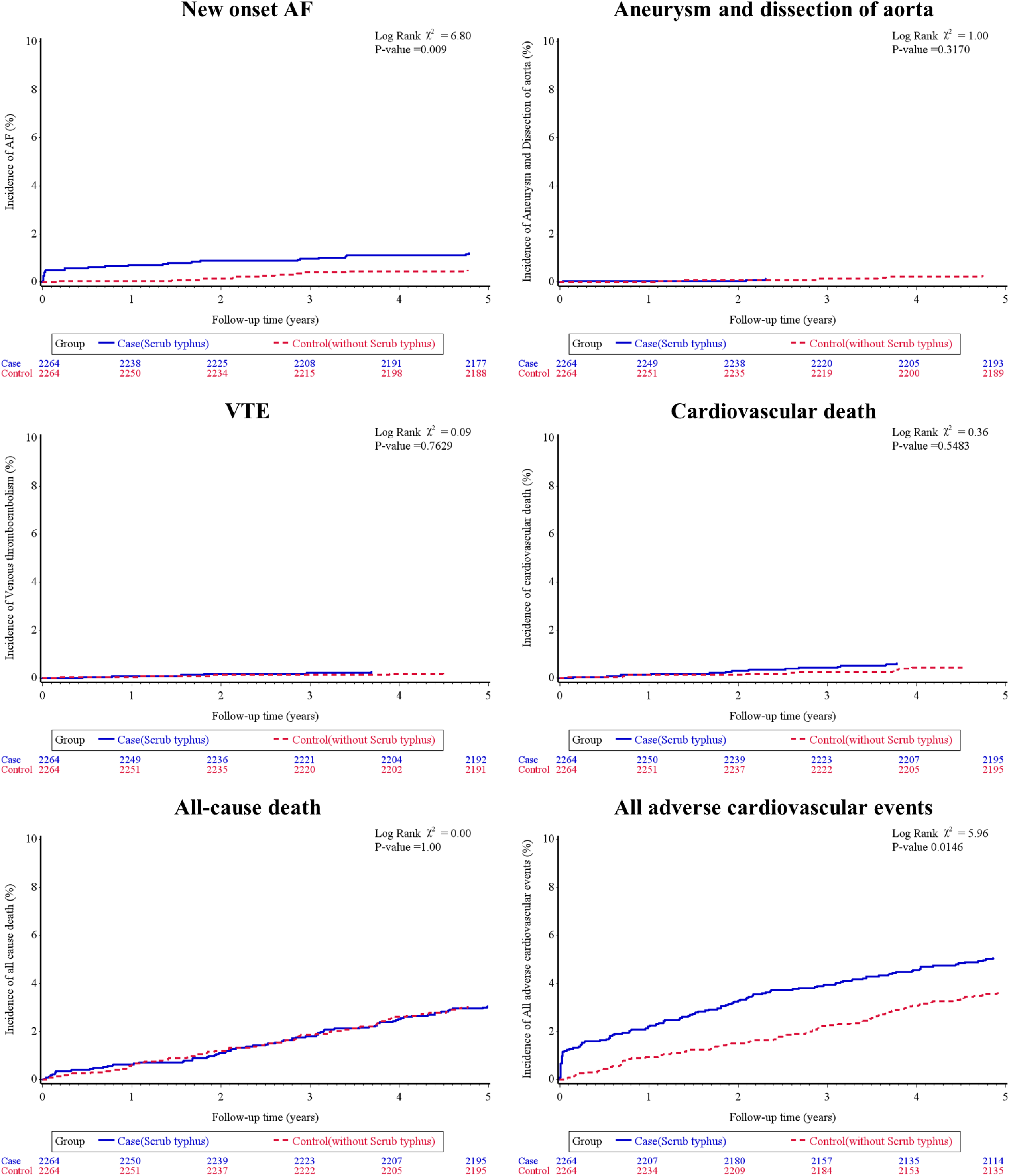
Cumulative incidence of outcome events of new-onset atrial fibrillation (AF) (A), aneurysm and dissection of aorta (B), venous thromboembolism (C), cardiovascular death (D), all-cause death (E), and all major cardiovascular events (F) in patients with and without scrub typhus.

**Table 1.**
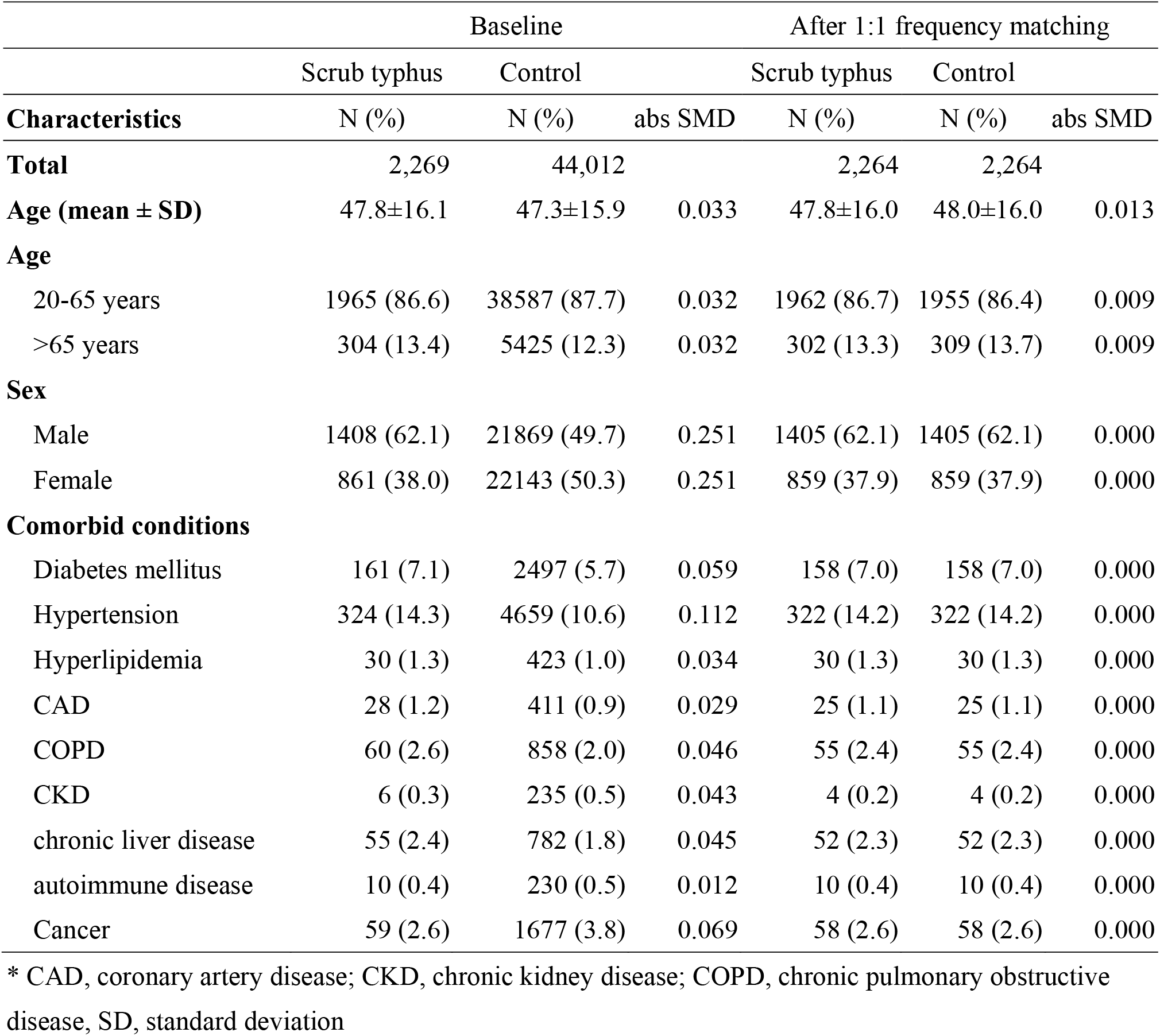
Baseline characteristics of study patients with Scrub typhus or without Scrub typhus.

|  | Baseline |  |  | After 1:1 frequency matching |  |  |
| --- | --- | --- | --- | --- | --- | --- |
|  | Scrub typhus | Control |  | Scrub typhus | Control |  |
| Characteristics | N (%) | N (%) | abs SMD | N (%) | N (%) | abs SMD |
| Total | 2,269 | 44,012 |  | 2,264 | 2,264 |  |
| Age (mean ± SD) | 47.8±16.1 | 47.3±15.9 | 0.033 | 47.8±16.0 | 48.0±16.0 | 0.013 |
| Age |  |  |  |  |  |  |
| 20-65 years | 1965 (86.6) | 38587 (87.7) | 0.032 | 1962 (86.7) | 1955 (86.4) | 0.009 |
| >65 years | 304 (13.4) | 5425 (12.3) | 0.032 | 302 (13.3) | 309 (13.7) | 0.009 |
| Sex |  |  |  |  |  |  |
| Male | 1408 (62.1) | 21869 (49.7) | 0.251 | 1405 (62.1) | 1405 (62.1) | 0.000 |
| Female | 861 (38.0) | 22143 (50.3) | 0.251 | 859 (37.9) | 859 (37.9) | 0.000 |
| Comorbid conditions |  |  |  |  |  |  |
| Diabetes mellitus | 161 (7.1) | 2497 (5.7) | 0.059 | 158 (7.0) | 158 (7.0) | 0.000 |
| Hypertension | 324 (14.3) | 4659 (10.6) | 0.112 | 322 (14.2) | 322 (14.2) | 0.000 |
| Hyperlipidemia | 30 (1.3) | 423 (1.0) | 0.034 | 30 (1.3) | 30 (1.3) | 0.000 |
| CAD | 28 (1.2) | 411 (0.9) | 0.029 | 25 (1.1) | 25 (1.1) | 0.000 |
| COPD | 60 (2.6) | 858 (2.0) | 0.046 | 55 (2.4) | 55 (2.4) | 0.000 |
| CKD | 6 (0.3) | 235 (0.5) | 0.043 | 4 (0.2) | 4 (0.2) | 0.000 |
| chronic liver disease | 55 (2.4) | 782 (1.8) | 0.045 | 52 (2.3) | 52 (2.3) | 0.000 |
| autoimmune disease | 10 (0.4) | 230 (0.5) | 0.012 | 10 (0.4) | 10 (0.4) | 0.000 |
| Cancer | 59 (2.6) | 1677 (3.8) | 0.069 | 58 (2.6) | 58 (2.6) | 0.000 |

\* CAD, coronary artery disease; CKD, chronic kidney disease; COPD, chronic pulmonary obstructive disease, SD, standard deviation

**Table 2.** Cardiovascular outcomes during 5-year follow-up.

|  | N | Events | Total<br>follow up | Incidence<br>rate (/1000<br>person-years) | Incidence rate<br>ratio (95% CI) | P value | Adjusted HR<br>(95% CI) | P value |
| --- | --- | --- | --- | --- | --- | --- | --- | --- |
| **AMI** |  |  |  |  |  |  |  |  |
| Case | 2264 | 25 | 10265.39 | 2.44 | 1.26 (0.70-2.27) | 0.441 | 1.26 (0.70-2.28) | 0.434 |
| Control | 2264 | 20 | 10346.89 | 1.93 |  |  |  |  |
| **HFH** |  |  |  |  |  |  |  |  |
| Case | 2264 | 37 | 10233.68 | 3.62 | 1.92 (1.10-3.34) | 0.022 | 1.97 (1.13-3.42) | 0.016 |
| Control | 2264 | 19 | 10350.28 | 1.84 |  |  |  |  |
| **Hemorrhagic stroke** |  |  |  |  |  |  |  |  |
| Case | 2264 | 13 | 10309.24 | 1.26 | 0.93 (0.44-1.99) | 0.860 | 0.94 (0.44-1.99) | 0.866 |
| Control | 2264 | 14 | 10374.29 | 1.35 |  |  |  |  |
| **Ischemic stroke** |  |  |  |  |  |  |  |  |
| Case | 2264 | 29 | 10275.01 | 2.82 | 0.86 (0.52-1.41) | 0.549 | 0.86 (0.53-1.42) | 0.557 |
| Control | 2264 | 34 | 10351.87 | 3.28 |  |  |  |  |
| **New onset AF** |  |  |  |  |  |  |  |  |
| Case | 2264 | 27 | 10264.46 | 2.63 | 2.39 (1.18-4.83) | 0.016 | 2.48 (1.23-5) | 0.011 |
| Control | 2264 | 11 | 10369.63 | 1.06 |  |  |  |  |
| **Aneurysm and**<br>**dissection of aorta** |  |  |  |  |  |  |  |  |
| Case | 2264 | 3 | 10308.48 | 0.29 | 0.50 (0.13-2.01) | 0.332 | 0.51 (0.13-2.02) | 0.334 |
| Control | 2264 | 6 | 10377.78 | 0.58 |  |  |  |  |
| **VTE** |  |  |  |  |  |  |  |  |
| Case | 2264 | 6 | 10305.39 | 0.58 | 1.21 (0.37-3.96) | 0.758 | 1.21 (0.37-3.97) | 0.751 |
| Control | 2264 | 5 | 10376.79 | 0.48 |  |  |  |  |
| **Cardiovascular death** |  |  |  |  |  |  |  |  |
| Case | 2264 | 14 | 10315.02 | 1.36 | 1.28 (0.58-2.82) | 0.539 | 1.28 (0.58-2.83) | 0.536 |
| Control | 2264 | 11 | 10381.49 | 1.06 |  |  |  |  |
| **All-cause death** |  |  |  |  |  |  |  |  |
| Case | 2264 | 69 | 10315.02 | 6.69 | 1.01 (0.72-1.41) | 0.970 | 1.01 (0.72-1.41) | 0.959 |
| Control | 2264 | 69 | 10381.49 | 6.65 |  |  |  |  |
| **All adverse**<br>**cardiovascular events** |  |  |  |  |  |  |  |  |
| Case | 2264 | 114 | 10110.61 | 11.28 | 1.41 (1.06-1.87) | 0.019 | 1.43 (1.08-1.91) | 0.013 |
| Control | 2264 | 81 | 10282.64 | 7.88 |  |  |  |  |

\*AF, atrial fibrillation; AMI, acute myocardial infarction; HFH, heart failure hospitalization; HR, hazard ratio; VTE, venous thromboembolism
\*All adverse cardiovascular events: AMI, HFH, hemorrhagic stroke, ischemic stroke, new onset atrial fibrillation, aneurysm and dissection of aorta, VTE, and cardiovascular death

**Table 3.**
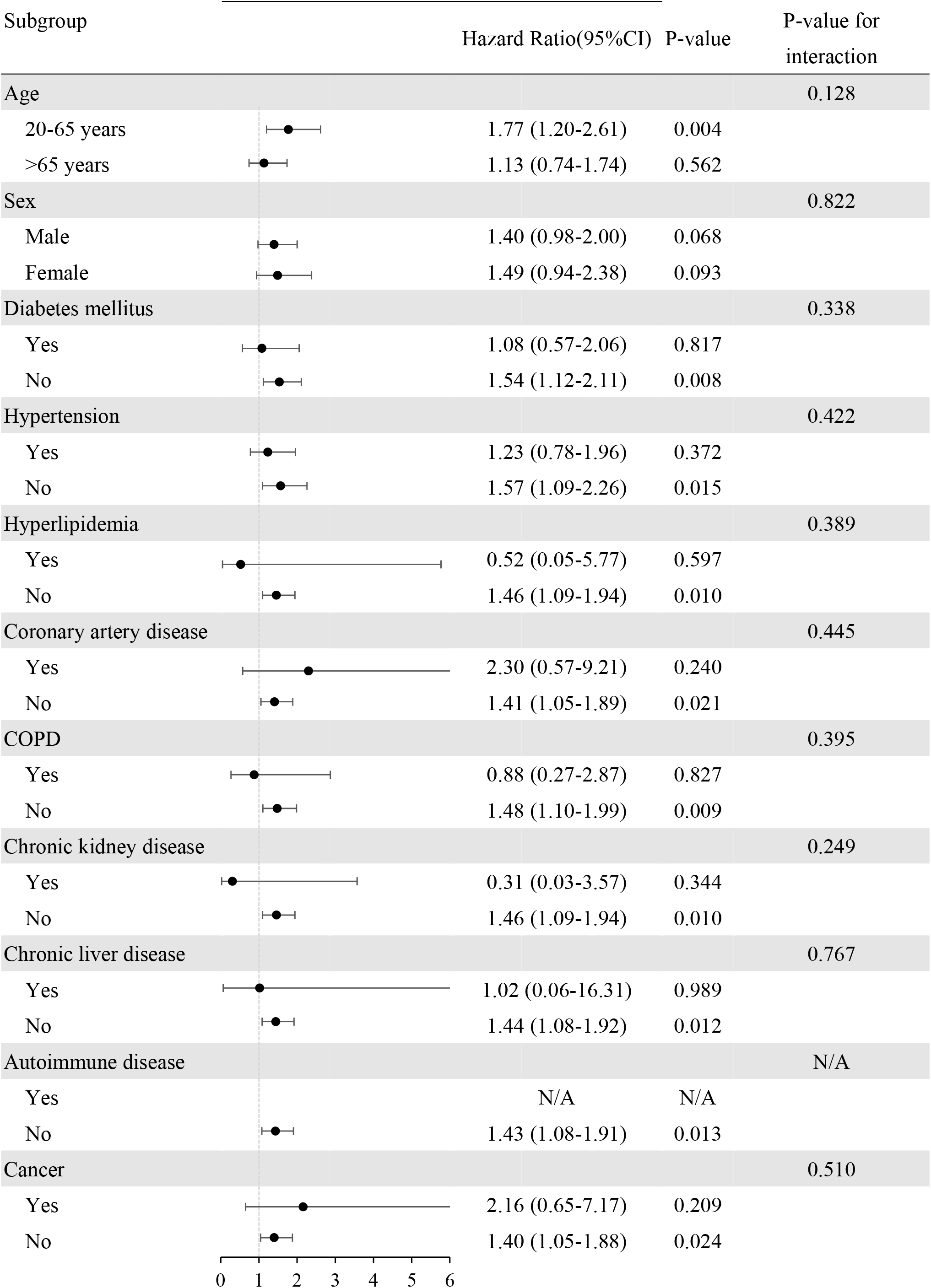
Subgroup analysis for the risk of all adverse cardiovascular events.

| Subgroup |  | Hazard Ratio(95%CI) | P-value | P-value for interaction |
| --- | --- | --- | --- | --- |
| Age |  |  |  | 0.128 |
| 20-65 years |  | 1.77 (1.20-2.61) | 0.004 |  |
| >65 years |  | 1.13 (0.74-1.74) | 0.562 |  |
| Sex |  |  |  | 0.822 |
| Male |  | 1.40 (0.98-2.00) | 0.068 |  |
| Female |  | 1.49 (0.94-2.38) | 0.093 |  |
| Diabetes mellitus |  |  |  | 0.338 |
| Yes |  | 1.08 (0.57-2.06) | 0.817 |  |
| No |  | 1.54 (1.12-2.11) | 0.008 |  |
| Hypertension |  |  |  | 0.422 |
| Yes |  | 1.23 (0.78-1.96) | 0.372 |  |
| No |  | 1.57 (1.09-2.26) | 0.015 |  |
| Hyperlipidemia |  |  |  | 0.389 |
| Yes |  | 0.52 (0.05-5.77) | 0.597 |  |
| No |  | 1.46 (1.09-1.94) | 0.010 |  |
| Coronary artery disease |  |  |  | 0.445 |
| Yes |  | 2.30 (0.57-9.21) | 0.240 |  |
| No |  | 1.41 (1.05-1.89) | 0.021 |  |
| COPD |  |  |  | 0.395 |
| Yes |  | 0.88 (0.27-2.87) | 0.827 |  |
| No |  | 1.48 (1.10-1.99) | 0.009 |  |
| Chronic kidney disease |  |  |  | 0.249 |
| Yes |  | 0.31 (0.03-3.57) | 0.344 |  |
| No |  | 1.46 (1.09-1.94) | 0.010 |  |
| Chronic liver disease |  |  |  | 0.767 |
| Yes |  | 1.02 (0.06-16.31) | 0.989 |  |
| No |  | 1.44 (1.08-1.92) | 0.012 |  |
| Autoimmune disease |  |  |  | N/A |
| Yes |  | N/A | N/A |  |
| No |  | 1.43 (1.08-1.91) | 0.013 |  |
| Cancer |  |  |  | 0.510 |
| Yes |  | 2.16 (0.65-7.17) | 0.209 |  |
| No |  | 1.40 (1.05-1.88) | 0.024 |  |

## Discussion

This is a nationwide longitudinal observational study to evaluate the long-term CV effect in patients infected with scrub typhus. First, we identified 1.97-fold and 2.48-fold increased incidence of hospitalization for heart failure and new-onset AF after patients infected with scrub typhus compared with matched population, respectively (aHR 1.97, 95% CI: 1.13-3.42; 2.48, 95%CI: 1.23-5.0). Second, patients with scrub typhus had a heighted risk for all adverse CV events during five-year follow-up after acute infection (aHR 1.43, 95% CI: 1.08-1.91). Third, the incidence of AMI, cerebrovascular accident, aortic disease, VTE and all-cause death did not differ between patients who had ever infected with or without scrub typhus in the cohort.

### Pathogenesis of Scrub Typhus

Current knowledge for molecular pathogenesis and associated immune responses of scrub typhus is still limited. From murine models of intradermal infection, O. tsutsugamushi escapes from autophagic elimination and replicates within mononuclear phagocytes and endothelial cells. It spreads from the skin inoculation site, regional lymph nodes to the multiple target organs and causes organ-specific pathologic features^13^. In murine models of severe infection, strong Th1/CD8-mediated immune responses, but suppressed Th2/regulatory T immunological markers, accompanied with loss of vascular endothelial integrity were discovered^14^. It is implicative of dysregulated and exacerbated immune responses, rather than direct injury of bacterial replications or cytotoxic effects, contributed to tissue damage, organ dysfunction and mortality^15^. However, important questions regarding to the critical elements triggering the host inappropriate immune responses and the time window to restore the balance and avoid serious complications still need to be investigated.

### Risks of Heart failure Hospitalization in Scrub Typhus

Heart failure syndrome due to acute scrub typhus infection may underestimate in previous literature^16^. In fact, two prospective cohort studies evaluate cardiac manifestations in scrub typhus with echocardiography and cardiac biomarkers, showing 30.9-42.8% patients had reduced ejection fraction; 61.7-72.8% patients had elevated levels of troponin T or creatine kinase-muscle/brain isoenzyme^9, 10^. The increased levels of cardiac biomarkers were correlated with the degree of systolic dysfunction. It indicated that cardiac injury and systolic dysfunction in acute scrub typhus infection are not uncommon complications. Although the immune response is activated to eliminate microbial-infected cells, the dysregulated inflammation causes excessive structural and functional abnormalities in cardiomyocytes with consequence of contractility failure, chamber remodeling, or conduction disturbances. Moreover, *O. tsutsugamushi* cause persistent or latent infection in some of patients who had recovered from acute illness^17^. Incubated macrophages remain continuously activated, which can hinder the resolution of inflammatory reactions and delay tissue-healing processes. In this cohort study, we indeed observed the increased risk of HFH in patients who was recovery and discharge from scrub typhus. To the best of our knowledge, this finding has not been previously reported in existing studies. This discovery carries two significant implications. First, early recognition and timely management with neurohormonal blockade are crucial for mitigating HF complications following scrub typhus infection. Evidence-based interventions for HF offer an opportunity to reverse cardiac pathological remodeling and enhance patient prognosis. Second, cardiac surveillance through echocardiography should be considered after an acute episode, particularly in the presence of associated symptoms and signs. Furthermore, shortening the time between symptom onset and accurate diagnosis could help eliminate pathogens before excessive inflammatory reactions and tissue destruction occur in the heart. Achieving this goal relies on increased disease awareness and the utilization of new diagnostic tools, which can confirm the diagnosis even when serologic tests yield negative results^18^.

### Risk of Atherosclerotic Cardiovascular Disease in Scrub Typhus

Chronic persistent inflammation is one of pathophysiologic mechanism of atherosclerotic cardiovascular diseases^19^. Microbial products activate innate immune system through the toll-like receptors and provoke a wave of inflammatory reactions within vascular atheroma^20^. *O.tsutsugamushi* in scrub typhus causes rickettsemia and replicates or harbors in the vascular endothelium and immune cells. It raises concern about the relationship between scrub typhus and atherosclerotic events. In a case series, transient ischemic attack and symptomatic CAD occurred 4-6 months after scrub typhus infection, with the persistence of viable *O. tsutsugamushi* confirmed through cell culture and nucleotide sequencing tool^17^. In a healthcare claims database study, patients with scrub typhus had 60% increased risk of subsequent AMI, but not cerebrovascular accident than control population^21^. Our results did not provide evidence supporting increased risks of developing ischemic stroke and AMI in patients with scrub typhus. The disparate results may arise from difference in selected patient cohort, prevalent prototype of bacterial strains, and follow-up duration between studies. Our patients who were young and survived from scrub typhus without prior CV events or serious complications, might be less likely to develop major atherosclerotic events in a shorter follow-up duration (4.5 versus over 10 years). Currently, it is still uncertain about the relationship between infectious disease and atherosclerotic event in the case of scrub typhus. Further prospective designed studies are needed to make an unequivocally conclusion.

### New Onset Atrial Fibrillation in scrub typhus

AF is the most common arrhythmia in the elderly and people with cardiovascular comorbidities. It often presents rapid and irregular electrical activities from multiple atrial foci. Current understanding of the mechanism of AF development mainly includes ectopic atrial activity in the muscular sleeves of the pulmonary vein ostia, structural and electrical property changes in the atria with multiple re-entrant circuits, and associated inflammatory process^22^. Inflammatory cascades cause atrial myocyte apoptosis, calcium overload, tissue fibrosis, and dysfunctional ion exchanger in sarcomeres, which contribute to initiation and perpetuation of AF^23, 24^. In our cohort, we observed an increased risk of new-onset atrial fibrillation (AF) in patients with scrub typhus. The presence of acute or chronic inflammation stimuli during scrub typhus can contribute to the development of "atrial myocarditis" and subsequent atrial remodeling. This association serves as the rationale for our findings. The importance and clinical perspective for AF occurrence in scrub typhus have been demonstrated in a previous cohort study^25^, which new-onset AF was found to be associated with three-month mortality and adverse cardiac complications, such as acute HF and ischemic heart disease. Moreover, paroxysmal AF identified in patient with scrub typhus predicted the confirmed diagnosis of acute myocarditis^26^. The nature course of acute myocarditis varied greatly among the recipients with time. Some patients with myocarditis presented with mild and self-limited symptoms, but other patients may develop refractory cardiogenic shock, requiring a mechanical circulation support. Early detection and proper management of scrub typhus associated myocarditis are paramount for prognosis and new-onset AF can be one of easily available and useful alarms to remind physician for possible severe cardiac complications.

### Study Limitation

Our study has several limitations. First, this is a retrospective, observational-designed study. Retrospective studies of specific infectious disease are usually limited to confounding by the characteristic of patients, which is vulnerable to infection or not. To address this, we included patients with scrub typhus who did not have prior cardiovascular events and matched them with a comparison cohort based on age, gender, and comorbidities. However, there may still be unmeasured biases between patients with and without scrub typhus. For instance, individuals infected with scrub typhus might tend to reside in rural areas with limited access to healthcare resources and lower awareness of the importance of cardiovascular disease prevention. Second, CV outcomes evaluated in the study were identified based on primary discharge diagnostic codes of hospitalization. The details of cardiac and laboratory exams, such as echocardiography and electrography, were not available in the claim-based database. The clinical manifestation of major CV diseases, like disease severity, cardiac chamber size, cardiac contractility, and arrhythmic burden, could not be evaluated and analyzed. Third, medications used for CV disease prevention, such as antihypertensive agents, antiarrhythmic drugs, or lipid-lowing agents were not included in the study. Although we did not have above information for statistical adjustment, individuals identified in the study were free from common CV events and less than one-tenth of people had diabetes mellitus, hyperlipidemia, CAD, or CKD. Nonetheless, this large-scale nationwide study with longitudinal follow-up for CV outcomes in patients with scrub typhus provides valuable clinically relevant information. Currently, there is a lack of high-level evidence in this field. Therefore, our results warrant well-designed prospective trials in the future to confirm the observations mentioned above.

## Conclusions

This nationwide retrospective cohort study identified an increased subsequent risk of CV events after scrub typhus infection. Patients with scrub typhus had higher rates of HFH and new-onset AF during the five-year follow-up. Inflammatory cascades triggered by the infectious agent can lead to irreversible CV system abnormalities. Although these abnormalities may be subclinical during the acute infection, they can contribute to CV events months or years later. This information highlights the need for physicians to be aware of the existing CV risk in these patients and consider essential cardiac surveillance when clinical suspicion is raised.

## Ethics approval and consent to participate

This study was conducted in accordance with the Declaration of Helsinki and the Declaration of Taipei on ethical considerations regarding health databases by the World Medical Association. The study protocol was approved, and informed consent was exempted by the Institutional Review Board of Chang Gung Medical Foundation (IRB No: 201901517B0) as all data converted from the original NHI’s claim records were anonymized and due to tight regulations of on-site analysis at HWDC.

## Consent for publication

Not applicable

## Availability of data and materials

All data generated or analyzed during this study are included in this published article.

## Data Availability

All data generated or analyzed during this study are included in this published article.

## Acknowledgements

The authors thank the statistical assistance and wish to acknowledge the support of the Maintenance Project of the Center for Big Data Analytics and Statistics Grant (CLRPG3D0049) at Chang Gung Memorial Hospital (CGMH) for study design and monitor, data analysis and interpretation. They also thank for the research grand support from the CGMH, Linkou (CMRPG3L0781) and the Ministry of Science and Technology (MoST 108-2410-H-182A-002) to Yu-Tung Huang. However, the funders had no role in study design, data collection and analysis, or preparation.

This study is based in part on data from the NID and NHIRD managed by Health and Welfare Data Science Center, Ministry of Health and Welfare (MoHW). However, the interpretation and conclusions contained herein do not represent the position of CGMH, NHI

Administration and MoHW.

## Conflict of interest

none

## Grants

Chang Gung Memorial Hospital Grant (CMRPG3L0781) and Ministry of Science and Technology Grant (MoST 108-2410-H-182A-002).

## Authors’ contributions

Study conception and design: JKY, VCW, SHC, YTH

Acquisition of data: SWC, CLW, YTH

Analysis and interpretation of data: JKY, VCW, YSL, CHC, MW,KCW

Drafting of manuscript: JKY, VCW

Critical revision: PHC, SHC, YTH

## Notes

### Competing Interest Statement

The authors have declared no competing interest.

## Reference

1 Costa, C. et al. Imported scrub typhus in Europe: report of three cases and a literature review. Travel Medicine and Infectious Disease 42, 102062 (2021).

2 Jiang, J. & Richards, A. L. Scrub typhus: no longer restricted to the Tsutsugamushi Triangle. Tropical medicine and infectious disease 3, 11 (2018).

3 Peter, J. V., Sudarsan, T. I., Prakash, J. A. J. & Varghese, G. M. Severe scrub typhus infection: Clinical features, diagnostic challenges and management. World journal of critical care medicine 4, 244 (2015).

4 Peter, J. V., Sudarsan, T. I., Prakash, J. A. & Varghese, G. M. Severe scrub typhus infection: Clinical features, diagnostic challenges and management. World J Crit Care Med 4, 244–250, doi:10.5492/wjccm.v4.i3.244 (2015).

5 Premraj, S. S., Mayilananthi, K., Krishnan, D., Padmanabhan, K. & Rajasekaran, D. Clinical profile and risk factors associated with severe scrub typhus infection among non-ICU patients in semi-urban south India. J Vector Borne Dis 55, 47–51, doi:10.4103/0972-9062.234626 (2018).

6 Varghese, G. M. et al. Scrub typhus in South India: clinical and laboratory manifestations, genetic variability, and outcome. Int J Infect Dis 17, e981–987, doi:10.1016/j.ijid.2013.05.017 (2013).

7 Thipmontree, W. et al. Scrub Typhus in Northeastern Thailand: Eschar Distribution, Abnormal Electrocardiographic Findings, and Predictors of Fatal Outcome. Am J Trop Med Hyg 95, 769–773, doi:10.4269/ajtmh.16-0088 (2016).

8 Hsu, Y. H. & Chen, H. I. Pulmonary pathology in patients associated with scrub typhus. Pathology 40, 268–271, doi:10.1080/00313020801911488 (2008).

9 Pannu, A. K. et al. Circulating cardiac biomarkers and echocardiographic abnormalities in patients with scrub typhus: A prospective cohort study from a tertiary care center in North India. J Vector Borne Dis 58, 193–198, doi:10.4103/0972-9062.321754 (2021).

10 Karthik, G. et al. Spectrum of cardiac manifestations and its relationship to outcomes in patients admitted with scrub typhus infection. World J Crit Care Med 7, 16–23, doi:10.5492/wjccm.v7.i1.16 (2018).

11 Choi, S. W. et al. Scrub Typhus and Abnormal Electrocardiography. Am J Trop Med Hyg 100, 399–404, doi:10.4269/ajtmh.17-0565 (2019).

12 Deluigi, C. C. et al. ECG findings in comparison to cardiovascular MR imaging in viral myocarditis. International Journal of Cardiology 165, 100–106, doi:https://doi.org/10.1016/j.ijcard.2011.07.090 (2013).

13 Keller, C. A. et al. Dissemination of Orientia tsutsugamushi and inflammatory responses in a murine model of scrub typhus. PLoS Negl Trop Dis 8, e3064, doi:10.1371/journal.pntd.0003064 (2014).

14 Shelite, T. R. et al. Hematogenously disseminated Orientia tsutsugamushi-infected murine model of scrub typhus [corrected]. PLoS Negl Trop Dis 8, e2966, doi:10.1371/journal.pntd.0002966 (2014).

15 Soong, L. Dysregulated Th1 Immune and Vascular Responses in Scrub Typhus Pathogenesis. J Immunol 200, 1233–1240, doi:10.4049/jimmunol.1701219 (2018).

16 Zhang, M. et al. Scrub typhus: surveillance, clinical profile and diagnostic issues in Shandong, China. Am J Trop Med Hyg 87, 1099–1104, doi:10.4269/ajtmh.2012.12-0306 (2012).

17 Chung, M. H., Lee, J. S., Baek, J. H., Kim, M. & Kang, J. S. Persistence of Orientia tsutsugamushi in humans. J Korean Med Sci 27, 231–235, doi:10.3346/jkms.2012.27.3.231 (2012).

18 Kannan, K. et al. Performance of molecular and serologic tests for the diagnosis of scrub typhus. PLoS Negl Trop Dis 14, e0008747, doi:10.1371/journal.pntd.0008747 (2020).

19 Henein, M. Y., Vancheri, S., Longo, G. & Vancheri, F. The Role of Inflammation in Cardiovascular Disease. Int J Mol Sci 23, doi:10.3390/ijms232112906 (2022).

20 Libby, P. et al. Inflammation, Immunity, and Infection in Atherothrombosis: JACC Review Topic of the Week. J Am Coll Cardiol 72, 2071–2081, doi:10.1016/j.jacc.2018.08.1043 (2018).

21 Jung, L. Y. et al. Risk of cerebro- and cardiovascular disease in patients with scrub typhus. Eur J Clin Microbiol Infect Dis 39, 451–454, doi:10.1007/s10096-019-03743-4 (2020).

22 Issac, T. T., Dokainish, H. & Lakkis, N. M. Role of inflammation in initiation and perpetuation of atrial fibrillation: a systematic review of the published data. J Am Coll Cardiol 50, 2021–2028, doi:10.1016/j.jacc.2007.06.054 (2007).

23 Aviles, R. J. et al. Inflammation as a risk factor for atrial fibrillation. Circulation 108, 3006–3010 (2003).

24 Frustaci, A. et al. Histological substrate of atrial biopsies in patients with lone atrial fibrillation. Circulation 96, 1180–1184 (1997).

25 Jang, S. Y. et al. New-onset atrial fibrillation predicting for complicating cardiac adverse outcome in scrub typhus infection. Clin Cardiol 42, 1210–1221, doi:10.1002/clc.23276 (2019).

26 Chin, J. Y., Kang, K. W., Moon, K. M., Kim, J. & Choi, Y. J. Predictors of acute myocarditis in complicated scrub typhus: an endemic province in the Republic of Korea. Korean J Intern Med 33, 323–330, doi:10.3904/kjim.2016.303 (2018).

